# Using Diffusion Similarity Networks to Identify, Track, and Predict Movement Disorders

**DOI:** 10.64898/2026.10.22.26366240

**Authors:** Ahmed Shalaby, Krishna Kanth Chitta, Amirreza Alijanpourotaghsara, Jeong Woo Choi, Frederick L. Hitti, Florence L. Chiang, Vibhash D. Sharma, Won Hwa Kim, Nader Pouratian, Jeon Lee

## Abstract

Characterizing individual structural brain alterations in movement disorders like Parkinson’s disease (PD) and Essential Tremor (ET) remains a significant challenge in clinical neuroscience. While brain structural connectivity (SC) derived from Diffusion Tensor Imaging (DTI) tractography maps physical white matter pathways, it fails to capture localized microstructural remodelings within gray matter structures. In this study, we use Diffusion Similarity Networks (DSNs) as a novel framework to estimate microstructural connectivity from diffusion weighted imaging (DWI). First, we evaluate the ability of DSNs to differentiate individuals with PD from healthy controls (HC). We used Similarity Network Fusion (SNF) to visualize differences between the fused networks of both groups as well as the Brain Connectivity Toolbox (BCT) to quantify changes in network topology, including transitivity, clustering coefficient, global efficiency, and betweenness centrality. Second, to evaluate the capacity of DSNs to track disease progression over time, we utilized longitudinal data extracted from the Parkinson’s Progression Markers Initiative (PPMI) database, mapping distinct functional subnetwork degeneration trajectories in PD patients. Finally, we developed a predictive Graph Neural Network (GNN) framework on a locally collected cohort to address the challenging task of differentiating PD from ET, utilizing DSNs as highly discriminant input features. Our results demonstrate that DSN metrics successfully capture distinct structural signatures that reliably separate clinical phenotypes, while longitudinal modeling reveals an accelerated, progressive decay in specific motor and basal ganglia subnetwork topologies over time. Furthermore, the GNN model achieved high classification performance (Balanced accuracy = 0.8684, AUC= 0.9420) in distinguishing PD from ET globally, with local explainability analysis successfully showing the specific brain Regions of Interest (ROIs) driving the differential diagnosis. Overall, these findings demonstrate that Diffusion Similarity Networks serve as a robust, non-invasive biomarker for movement disorders, offering deep insights into disease progression and an explainable AI tool for clinical decision support.

## 1 Introduction

Movement disorders cover a wide range of neurological conditions that disrupt how the body controls everyday movement. Diagnosing these conditions early in the disease course can be difficult because the initial changes in the brain are not detectable on standard clinical imaging, and initial symptom presentations can be mild and/or ambiguous [1]. Predicting how a patient’s disease will progress over time is also challenging, because two patients starting with the exact same symptoms can follow completely different paths. An example of this challenge is the common clinical overlap between Parkinson’s Disease (PD) and Essential Tremor (ET)[2]. In its advanced stages, PD involves midbrain degeneration accompanied by the hallmark motor symptoms of rigidity, resting tremor, and bradykinesia. Furthermore, PD patients often experience significant non-motor dysfunction such as sleep disturbances, pain, and cognitive impairment. On the other hand, ET involves mild loss of purkinje cells in the cerebellum and is primarily associated with action tremors. However, in the early stages of these diseases, both can present similarly with tremor. This diagnostic uncertainty can lead to misdiagnosis rates of 20% to 30% even at specialized clinics [3–5]. Because these two diseases require completely different treatments and have very different long-term outcomes, developing an objective biomarker that can accurately differentiate these diseases early in their disease courses and track their progression over time represents an urgent unmet need.

Imaging biomarkers are a compelling option because they can be obtained non-invasively in routine clinical practice. Magnetic resonance imaging (MRI), for example, can provide a detailed structural view of the brain and may be applied to study movement disorders [6, 7]. Among advanced imaging techniques, Diffusion Tensor Imaging (DTI) has demonstrated promise by modeling the directional diffusion of water molecules along axonal pathways, thereby providing a window into the microstructural white matter integrity [8]. Large-scale multi-site investigations have demonstrated that microstructural white matter alterations follow stage-specific trajectories in neurodegenerative conditions, altering fractional anisotropy (FA) and mean diffusivity (MD) profiles as tissue breakdown accelerates [9]. In PD individuals, localized DTI analysis has consistently identified structural deficits within the substantia nigra, internal capsule, cerebellar peduncles, and corticospinal tracts [10–13]. In contrast, ET individuals typically exhibit alterations within the cerebellothalamic and frontocerebellar circuits, supporting the hypothesis of an underlying cerebellar network dysfunction [14–16].

Neurodegenerative diseases do not simply damage single, isolated areas of the brain; instead, they disrupt the communication pathways across the entire brain network. Because of this, neuroimaging has shifted away from relatively simplistic morphometric analyses toward analyzing the brain as a whole interconnected system, a framework known as connectomics [17–19]. We can map this complex wiring diagram by capturing millions of white matter pathways into a structural connectivity (SC) matrix via tractography using diffusion-weighted MRI (dMRI) [18]. Viewing the brain through this perspective allows us to use graph theory to calculate how efficiently information flows through the system. Ultimately, these whole-brain mathematical wiring maps can be fed into machine learning models to automatically detect widespread patterns, making it possible for computers to accurately identify and classify different clinical disorders [19].

Recent breakthroughs in artificial intelligence, particularly deep learning, have revolutionized the analysis of high-dimensional neuroimaging graphs. Unlike traditional machine learning frameworks that depend on manual, potentially biased feature engineering, deep architectures such as Convolutional Neural Networks (CNNs) and Graph Neural Networks (GNNs) automatically learn hierarchical, non-linear representations directly from raw graph embeddings [20–22]. In movement disorders, frameworks utilizing Graph Attention Networks (GATs) have demonstrated high performance in separating PD subjects from healthy controls by highlighting highly informative connectivity signatures within subcortical loops [18]. Integrating deep learning with connectomics offers major advantages, including automated feature extraction from vast search spaces, the discovery of subtle and diffuse network configurations that escape standard statistical testing, and excellent generalizability when paired with rigorous feature selection protocols [23].

Despite these computational advances, traditional white matter SC frameworks suffer from fundamental limitations that restrict their clinical utility, especially in early-stage disease [24]. The SC matrices derived from traditional deterministic or probabilistic tractography strictly quantify physical white matter microstructure (e.g., fiber counts or streamline densities). Therefore, they fail to capture the subtle, higher-order, localized microstructural remodeling that occurs within gray matter structures and at the grey-white matter interface during early neurodegeneration [25]. In particular, structural network mapping approaches are shifting from traditional group-level structural covariance models toward single-scan architectonic and similarity networks [19]. These single-subject frameworks allow for the mapping of an individual’s unique connectome and open new avenues for precise phenotypic tracking [19]. In addition to diffusion imaging techniques, other advanced MR imaging modalities such as high-resolution susceptibility map-weighted imaging (SMwI) have been investigated as diagnostic tools to differentiate PD from ET. A recent study used SMwI to visualize the loss of dorsal nigral hyperintensity within the nigrosome-1 region, and demonstrated remarkable sensitivity and specificity in separating tremor-dominant PD from ET [26]. By isolating these microstructural and subcortical markers, researchers have successfully begun mapping invariant common core subnetwork signatures in PD that remain robust across different clinical phenotypes and advance predictably with disease duration [27].

While recent work has been promising, connectomic measures have not yet been used to differentiate between PD and ET. This study introduces an innovative, multi-stage framework centered on *Diffusion Similarity Networks (DSNs)* to estimate microstructural connectivity directly from DTI data [24]. Rather than tracking physical fiber pathways, DSNs utilize the statistical similarity of diffusion profiles between parcellated anatomical regions, making them highly sensitive to localized microstructural alterations within gray and white matter structures alike. Furthermore, to address the challenge of detecting parallel network decay over time, we introduce the *Combined Similarity Score (CSS)* metric. This new metric reduces the connection strength between any two brain regions if the individual regions themselves demonstrate tissue degeneration. Here, we apply these analyses to generate classifiers that reliably differentiate between ET and PD.

## 2 Motivations and Main Contributions

Sadikov et al. [24] established DSNs as a novel representation of gray-matter organization in healthy populations. Unlike conventional white-matter tractography, which maps physical axonal fiber pathways between regions [28], DSNs measure the architectural and microstructural homogeneity of gray matter itself [24, 29]. By modeling brain regions as similarity nodes, DSNs construct edge weights based on the statistical equivalence of inter-regional diffusion metrics (such as fractional anisotropy, mean diffusivity, or neurite density profiles) [24]. Physically, a high DSN edge weight indicates that two distinct cortical or subcortical areas share a closely matched intrinsic microstructural fingerprint—reflecting similar cellular packing densities, dendritic arborization, and local myelination patterns [24, 29]. This paradigm offers several critical advantages over traditional structural connectomics. First, DSNs provide direct access to gray-matter tissue health, bypassing the long-standing limitation of tractography in deep subcortical structures where fiber-crossing, high iron deposition, and complex terminal branching often cause severe tracking errors or signal loss [28, 30]. Second, DSNs capture the subtle microstructural degradation, such as early neuronal loss or neuroinflammation, prior to gross physical white-matter bundle breakdown [24, 29]. Consequently, DSNs offer an individualized, biologically interpretable representation of gray-matter structural organization that is orthogonal to conventional SC, making them uniquely suited for tracking diffuse neurodegenerative processes [24].

In this study, we demonstrate that DSNs can capture disease-relevant biomarkers, longitudinal neurodegeneration, and disease differentiation that have not previously been shown. First, we evaluated the fundamental sensitivity of the DSN framework pairing it with the similarity network fusion (SNF)[31] and the Brain Connectivity Toolbox (BCT)[32] to extract and compare global integration and subnetwork features. Second, we developed a Longitudinal Velocity Coupling framework that maps the brain similarity networks and clinical motor trajectories into a unified 2D coordinate space. This approach tracks the relationship between the velocity of loss of the microstructural network and the accumulation rate of the Movement Disorder Society-Unified Parkinson’s Disease Rating Scale Part III (MDS-UPDRS III) total motor score [33] over a four-year period. Additionally, we implemented a *K*-NN clustering approach utilizing the clinical score trajectories to stratify patients into clinically stable versus rapid progression phenotypes across distinct subnetwork tracks. Finally, we integrated these microstructural networks with an Iterative Permutation Sampling Recursive Feature Elimination (IPS-RFE) algorithm [34] and a predictive Graph Neural Network (GNN) architecture on a locally collected cohort to perform a differential classification between PD and ET using the underlying DSN topologies.

## 3 Materials and Methods

As shown in Fig 1, the workflow begins by constructing Diffusion Similarity Networks (DSNs) from raw DWI scans of participants. After preprocessing [35] the diffusion weighted images, the brain is split into 90 distinct regions based on the third version of the automated anatomical-labelling (AAL3)[36] anatomical atlas to build the DSNs, where each region is a node and the connections represent microstructural similarity. From there, the framework splits into three main applications to test how useful these networks are. First, to see if DSNs can differentiate between PD and HC, we fuse the networks using SNF [31] to visualize them and use the BCT [32] to calculate structural changes like efficiency and centrality. Second, we use longitudinal data from the PPMI dataset[37] to track how PD progresses over time, mapping out how specific brain subnetworks—like the motor and basal ganglia systems—degenerate. Finally, we feed these DSNs into a Graph Neural Network (GNN) using a local patient cohort to see how accurately the model can differentiate between PD and ET.

**Fig. 1.**
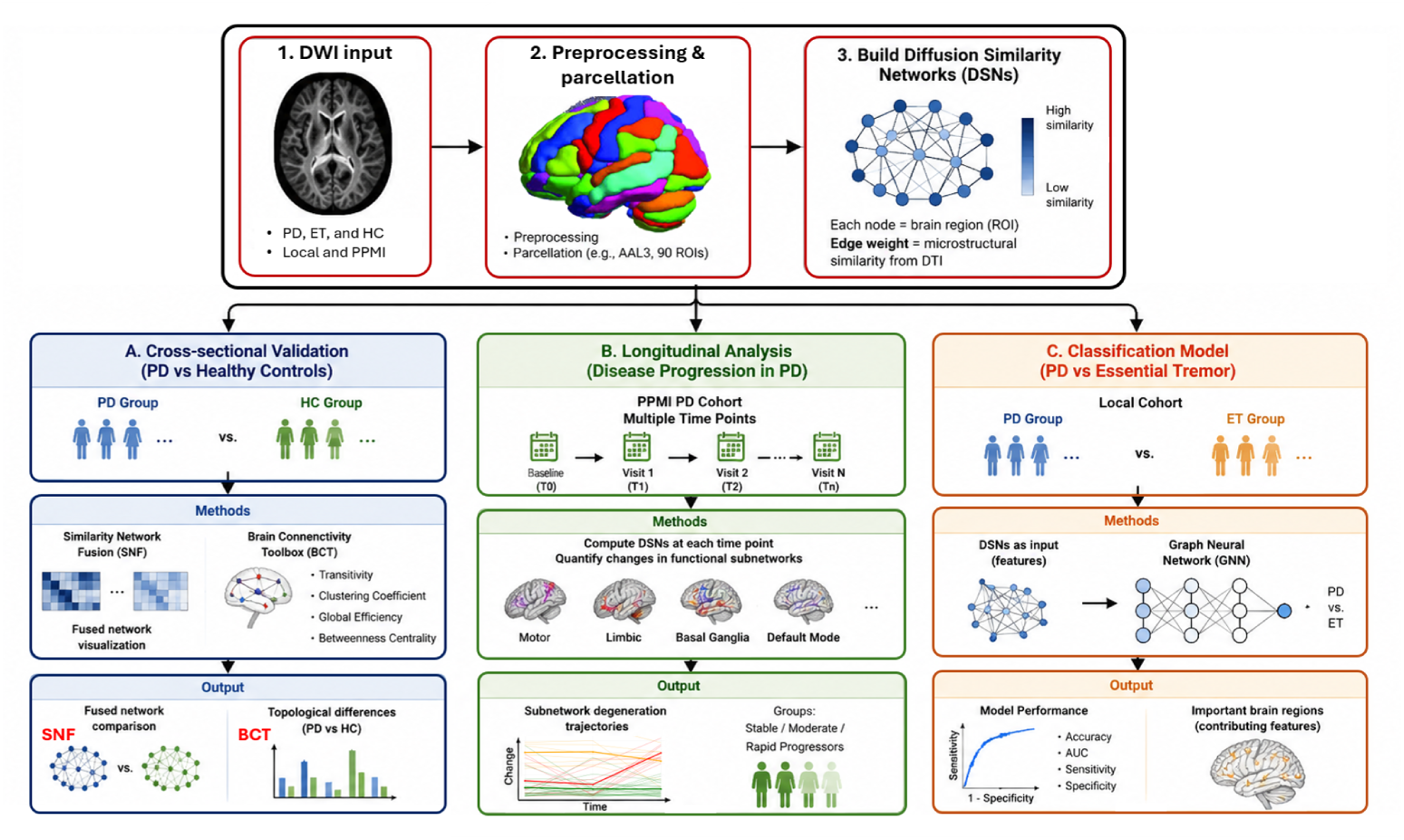
Overview of the study workflow. Diffusion imaging data were used to construct Diffusion Similarity Networks (DSNs), which were applied in three analyses: (1) cross-sectional comparison of PD and healthy controls, chosen from PPMI database (2) longitudinal analysis of disease progression using the same PPMI subset, and (3) Graph Neural Network (GNN)-based classification of PD and ET.

### 3.1 Dataset Description

Our neuroimaging and clinical datasets were obtained from two distinct sources. The first data source was the PPMI database [37]. To ensure robust cross-sectional comparisons and reliable longitudinal modeling, we selected a suitable sub-cohort consisting of 107 subjects, comprising 41 individuals diagnosed with PD and 66 HC. These two groups were carefully matched for age and sex to minimize confounding structural variations. All subjects were required to have comprehensive clinical characterization, specifically MDS-UPDRS III total motor scores [33]. For the longitudinal analysis, the 41 patients with PD obtained a baseline scan (T0) and three subsequent follow-up scans acquired approximately 1 year (T1), 2 years (T2) and 4 years (T3) after the baseline scan.

The second dataset is comprised of a locally collected cohort (obtained at UT Southwestern medical center) and included 100 individuals, 57 diagnosed with PD and 43 with ET. All individuals underwent whole-brain MRI on a 3T Philips Ingenia scanner. In addition to pre- and post-contrast T1-weighted MRI (T1w), diffusion-weighted MRI (dMRI) was acquired using a single-shell spin-echo EPI sequence with 32 diffusion-encoding directions (b = 800 s/mm²) and two opposite phase-encoding directions with an isotropic resolution of 2.5 mm. Each acquisition included one *b*_0_ image, resulting in 33 volumes per phase-encoding direction [38]. Data were collected with the approval of the Institutional Review Board (IRB) number: STU-2024-0337. All methods were performed in accordance with the relevant guidelines and the Health Insurance Portability and Accountability Act (HIPAA) regulations. All participants provided their informed written consent in accordance with the Declaration of Helsinki. The raw DWI data were corrected using the standard FMRIB software library (FSL) preprocessing pipeline [35], including motion and eddy-current correction. The participants’ demographics are presented in Table 1. The mean age was 62.9 ± 11.6 years, and approximately 75% of individuals were male.

**Table 1.** Participants’ demographic characteristics of the UTSW dataset.

| Characteristic | Parkinson's Disease | Essential Tremor |
| --- | --- | --- |
| Number of individuals | 57 | 43 |
| Mean age (years) | $61.8 \pm 11.2$ | $64.4 \pm 12.1$ |
| Sex (male) | 46 (80.7%) | 29 (67.4%) |
| Imaging status | Pre-surgical (DBS) | Pre-surgical (DBS) |

### 3.2 DWI Preprocessing and Brain Parcellation

DWI data were preprocessed using FSL pipeline (version 6.0.7.14) [35]. Distortions due to head motion and eddy currents were corrected via affine registration implemented in FMRIB’s linear image registration tool (FLIRT). Brain extraction was subsequently performed on the DWI images using the FSL Brain Extraction Tool (BET). Finally, the diffusion tensors and voxelwise diffusion parameters were estimated using DTIFIT implemented in FSl pipeline.

Each individual’s diffusion data were parcellated into 90 cortical and subcortical regions using the AAL3 atlas (Figure 2) The brain parcellation was performed using a series of structural and diffusion image preprocessing and registration steps. Structural T1w images were first skull-stripped using the FSL BET. The resulting brain-extracted T1 images were then affinely registered to the 2 mm resolution Montreal Neurological Institute (MNI) standard space using FSL FLIRT with 12 degrees of freedom. To ensure accurate alignment between modalities, the skull-stripped T1, and DWI images of each individual were then coregistered using a rigid-body transformation with six degrees of freedom, implemented via the FSL with a boundary-based registration (BBR) cost function, which has been shown to outperform single-stage registration approaches. The transformation matrices from these steps were combined and applied to warp the AAL3 atlas ROI masks from MNI space into each individual’s native DWI space.

**Fig. 2.**
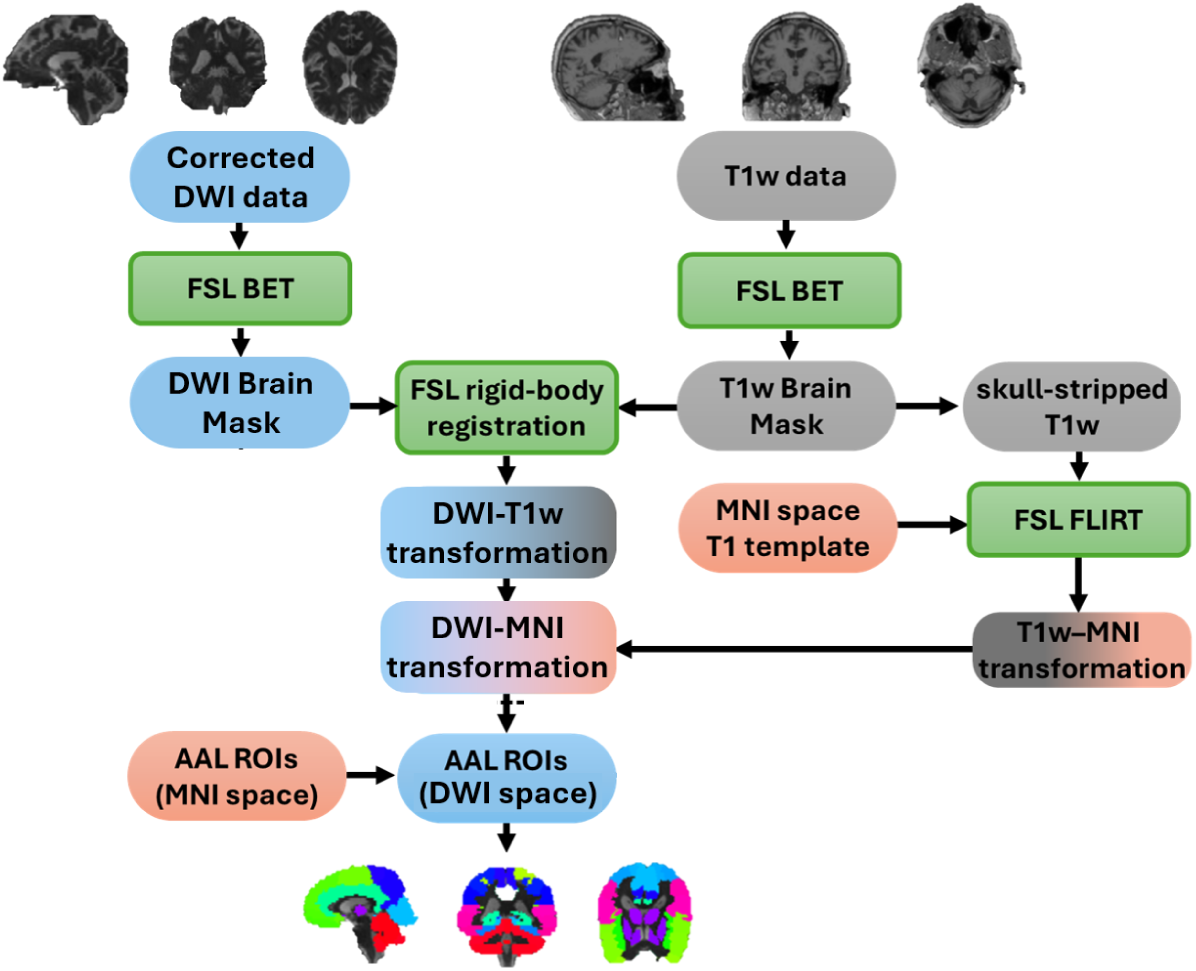
Brain parcellation steps using FSL pipeline and the AAL3 atals.

### 3.3 Estimation of Diffusion Similarity Network (DSN)

As shown in Fig. 3, the estimation of individual-specific DSNs follows a multi-stage voxel-wise density configuration that avoids reliance on white matter tractography [24]. Given a spherically sampled diffusion MRI shell, the raw diffusion-weighted signal *f* (*θ, φ*) inside each voxel is linearly decomposed onto an orthonormal spherical harmonic basis:

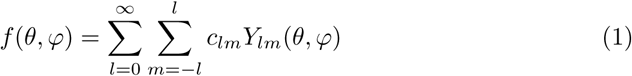

**Fig. 3.**
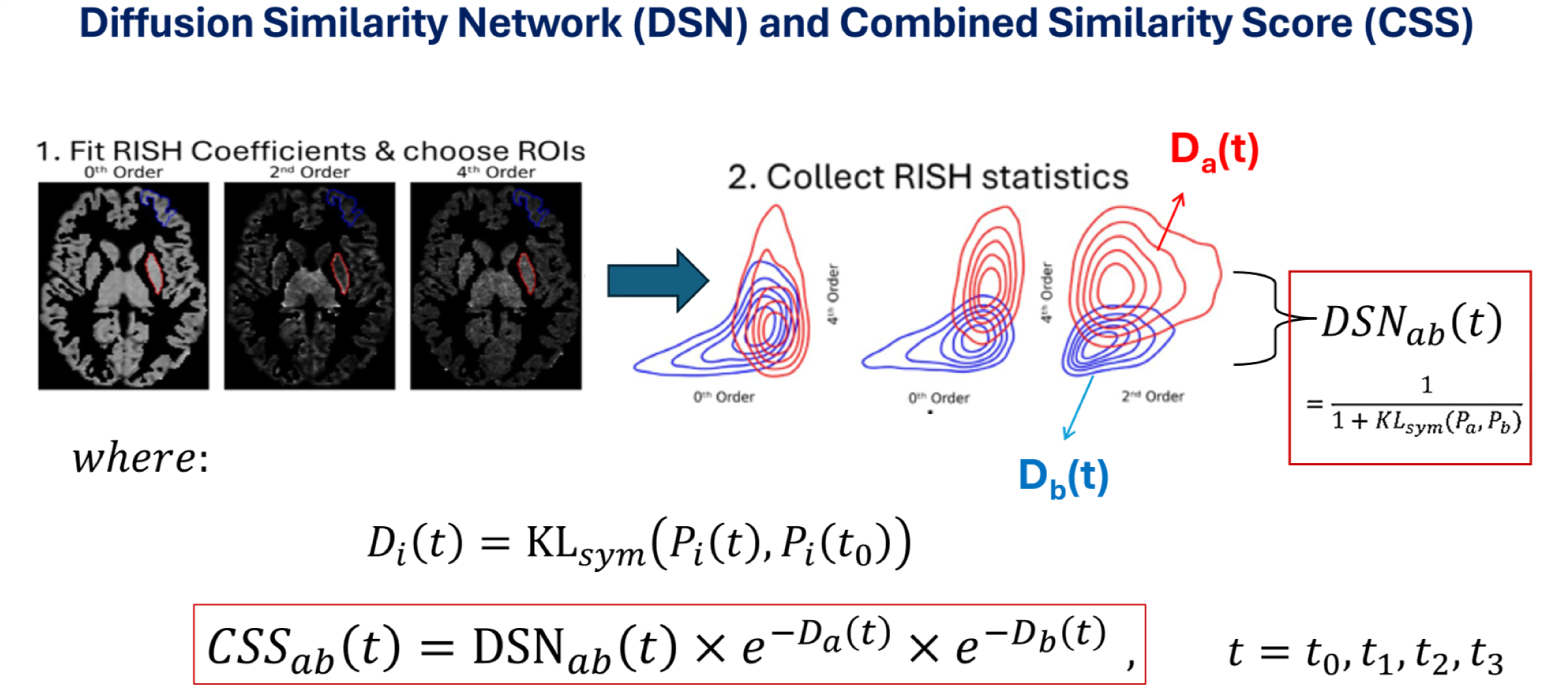
DSN estimation and CSS modification.

To ensure strict rotational invariance, the total energy within each individual frequency profile *l* is extracted across even degrees (*l* = 0, 2, 4) to match diffusion symmetry constraints, generating the rotationally invariant spherical harmonic (RISH) descriptors:

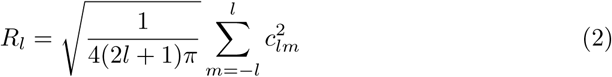

For each subject, these RISH values are generated across voxels encapsulated within the specific anatomical boundaries dictated by a chosen parcellation scheme. Letting *P_a_* and *P_b_* denote the multi-dimensional empirical probability distributions of RISH vectors collected across all voxels spanning ROI *a* and ROI *b* respectively, the structural edge connectivity weight defining the mutual relationship matrix is obtained via the inverse of the symmetrized Kullback-Leibler (KL) divergence [39] using a *k*-nearest neighbors estimator:

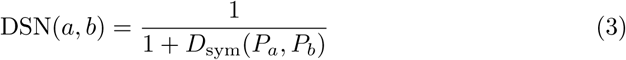

where *D*_sym_(*P_a_, P_b_*) = *D*_KL_(*P_a_||P_b_*) + *D*_KL_(*P_b_||P_a_*). Throughout this study, network node specifications are mapped according to AAL3 template, ultimately yielding a dense, symmetric 90 *×* 90 gray matter microstructural similarity matrix for every individual scan.

While the standard DSN effectively captures the microstructural similarity between two regions of interest (ROIs) at a static time point, it introduces a significant limitation when applied to longitudinal data. If two distinct brain regions (*ROI_A_* and *ROI_B_*) undergo a simultaneous and identical rate of microstructural degradation over time, their relative DTI metric profiles will remain highly correlated. Consequently, the standard DSN will yield a high similarity score at later time points, failing to reflect the actual physical degeneration occurring within each individual region.

To overcome this limitation and accurately capture absolute longitudinal changes, we developed the Combined Similarity Score (CSS) metric. The CSS modulates the baseline DSN value by accounting for the independent microstructural integrity of each region, ensuring that absolute tissue degradation penalizes the final connectivity weight. For any two regions *a* and *b* at a given time point, the CSS is mathematically defined as:

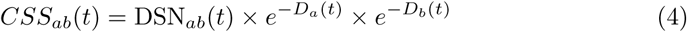

where DSN*_ab_* represents the standard diffusion similarity between the two regions, while *D_a_* and *D_b_* denote the quantified structural degradation or deviation metrics calculated for *ROI_a_* and *ROI_b_* relative to the baseline scan, respectively. By integrating these exponential decay terms, CSS ensures that even if two regions have highly correlated profiles, their net connectivity weight will systematically decrease if the underlying tissues are actively degenerating. Due to its sensitivity to parallel microstructural decline, this CSS framework is explicitly deployed in the longitudinal experiment of our study to map and track functional subnetwork degeneration trajectories in PD over time.

### 3.4 Longitudinal Velocity Coupling Framework

To establish a direct relation between microstructural brain network decay and clinical motor decline, we introduced a novel Longitudinal Velocity Coupling framework (Figure 4). For each individual patient, we mapped two parallel trajectories across the 4-year period: (1) the microstructural trajectory, derived from the change in CSS values across sessions, and (2) the clinical trajectory, derived from the corresponding longitudinal MDS-UPDRS Part III motor scores.

**Fig. 4.**
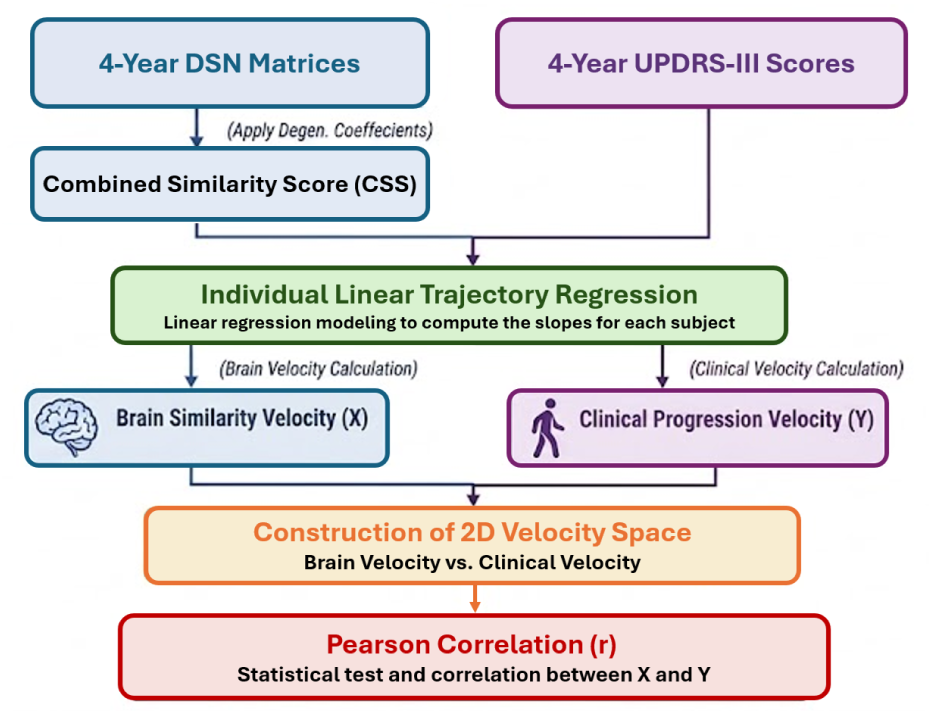
The pipeline of the Longitudinal Velocity Coupling framework. For each patient, linear regression trajectories are fitted across four time points to calculate the Brain Similarity Velocity (*X*, rate of CSS loss) and the Clinical Progression Velocity (*Y*, rate of MDS-UPDRS III accumulation). Pearson correlation analysis is then performed in the combined 2D coordinate space to determine if structural network decay drives clinical decline.

An individual ordinary least-squares (OLS) linear regression model was fitted to each trajectory to compute its slope over time [40, 41]. This process yields two primary metrics per subject: the *Brain Similarity Velocity* (*X*), representing the individual rate of microstructural connectivity loss, and the *Clinical Progression Velocity* (*Y*), representing the individual rate of clinical motor symptom accumulation. Every patient is thus compressed into a single coordinate point within a unified 2D velocity space. Finally, a Pearson correlation coefficient (*r*) was calculated across the cohort to assess the relationship between fast microstructural network decay and accelerated clinical decline.

### 3.5 Feature Selection Using IPS-RFE and GNN Classification

To generate a diagnostic classifier, each individual’s brain network was represented as a graph *G* = (*V, E,* Ω), where *V* denotes the set of nodes corresponding to the 90 parcellated brain regions, *E* represents the edges, and Ω is the symmetric 90 *×* 90 edge weight (the element of the DSN matrix). The upper triangle of this matrix yields a high-dimensional feature space of 4,005 unique microstructural similarity connections (90*×*89*/*2). Given the cohort size of 100 individuals, this introduces a pronounced high-dimensional learning challenge. To reduce noise and isolate the most discriminative connections prior to graph construction, we utilized our previously published Iterative Partial Scoring Recursive Feature Elimination (IPS-RFE) framework [34]. Within this framework, a *Random Forest (RF)* classifier [42] served as the base estimator, ranking feature importance via the *GINI impurity* (*IG*), defined as:

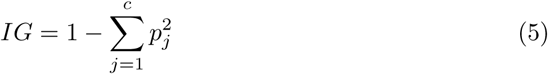

where *p_j_* denotes the probability of class *j* among the *c* classes. To prevent overfitting and ensure robust generalization, the RFE process was integrated with an *iterative permutation sampling (IPS)* strategy. At each iteration, a fraction *f* = 0.8 of the dataset was randomly sampled, and 10-fold cross-validation was performed. Connections were accumulated and ranked based on their selection frequency across iterations, and the top 15% (601 connections) were retained to form the final reduced DSN feature space.

Following feature selection, the filtered DSN was modeled as an *undirected weighted graph* and fed into a Graph Neural Network (GNN) for graph-level classification. To effectively integrate edge weights during node representation updates, we employed two *GraphConv* layers to aggregate localized neighbor profiles while minimizing over-smoothing. The node update operation is expressed as:

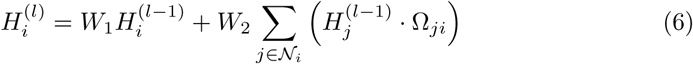

where *W*_1_ and *W*_2_ are learnable parameter matrices, 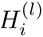 represents the feature vector of node *v_i_* at layer *l*, *N_i_* is the set of neighboring nodes, and Ω*_ji_* is the DSN edge weight between nodes *v_j_* and *v_i_*. Batch normalization layers were applied after each graph convolution to ensure training stability. Subsequently, a *global max pooling* layer was used to aggregate node embeddings into a single graph-level representation:

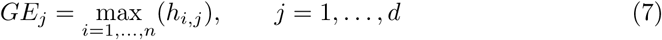

where *h_i,j_* represents the *j*^th^ feature of the *i*^th^ node, *n* = 90 is the total number of nodes, *d* = 64 is the embedding vector length, and *GE_j_*denotes the final graph embedding element. This unified representation was vectorized and passed through a multilayer perceptron (MLP) to output the final diagnostic label (PD or ET). The architectural details and structural dimensionality transitions of the network are detailed in Table 2. The pipeline of the proposed system is illustrated in Fig. 5.

**Fig. 5.**
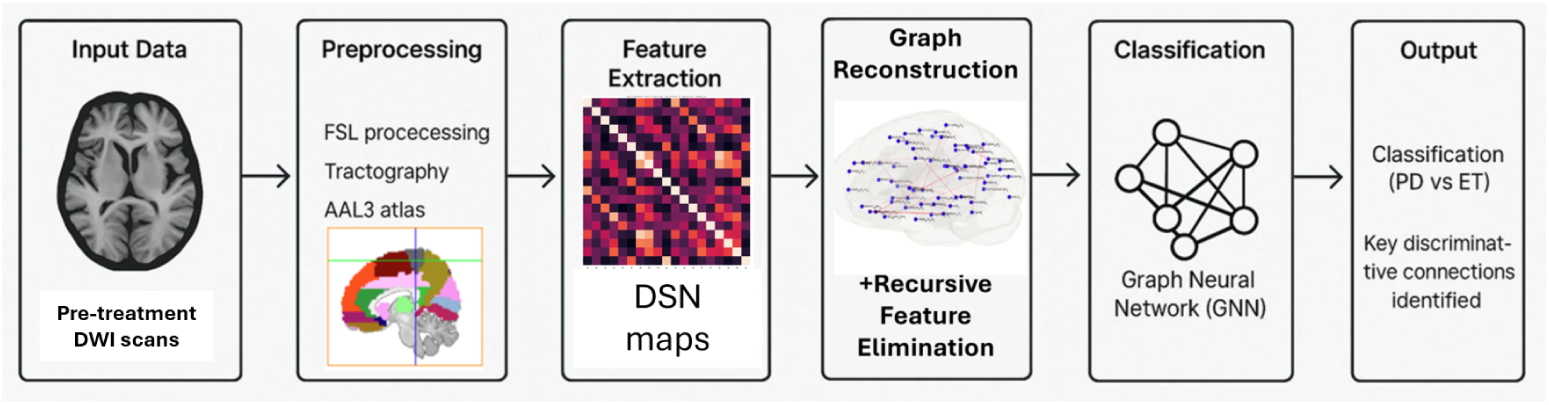
The framework of the proposed IPS-RFE GNN model for PD vs. ET classification.

**Table 2.** The GNN Model Architecture and Data Dimensions using DSN Features.

| Stage | Input Shape | Operation | Output Shape |
| --- | --- | --- | --- |
| Raw Input | $90 \times 90$ | Load Individual DSN Matrix | $[90 \times 90]$ |
| RFE Pre-processing | Vector 4005 | Flatten Upper Triangle | Vector 4005 |
| Feature Selection | Vector 4005 | Random Forest (Top 601) | Vector 601 |
| GNN Input ($H^0$) | $90 \times 90$ | Reconstruction | $[90 \times 90]$ |
| GraphConv Layer 1 | $[90 \times 90]$ | GraphConv(90, 64) + ReLU | $[90 \times 64]$ |
| GraphConv Layer 2 | $[90 \times 64]$ | GraphConv(64, 64) + ReLU | $[90 \times 64]$ |
| Readout Layer | $[90 \times 64]$ | Global Max Pooling | $[1 \times 64]$ |
| FC Hidden Layer | $[1 \times 64]$ | Linear (64, 32) + ReLU | $[1 \times 32]$ |
| Classification | $[1 \times 32]$ | Linear (32, 2) | $[1 \times 2]$ |
| Prediction | $[1 \times 2]$ | Softmax / Argmax | Scalar |

## 4 Experimental Results

As a proof of concept and to empirically validate our main hypothesis that DSNs capture intrinsic gray-matter (GM) microstructural organization orthogonal to white-matter structural connectivity (SC), we first evaluated the regional and subject-level coupling between brain network topologies and GM morphology (cortical thickness) for the HC group.

Mathematically, for each subject *k ∈ {*1*, …, M}*, the individual DSN matrix **D**^(^*^k^*^)^ *∈* ℝ*^N×N^* is constructed by computing the pairwise statistical similarity of microstructural diffusion features across *N* parcellated regions, setting self-loop diagonals 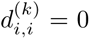. The nodal strength 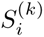 for region *i* is quantified as the total sum of its weighted similarity edges:

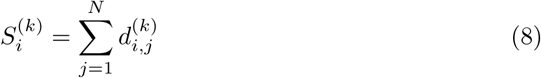

The regional nodal strength averaged by the population is given by 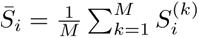. We evaluated biological alignment by computing the Pearson correlation coefficient (*r*) between 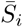 and the mean cortical thickness of the cohort 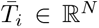, comparing DSN with the SC nodal strengths derived from conventional tractography under identical conditions. Statistical significance was evaluated using Steiger’s *Z*-test [43] for dependent correlations.

DSN nodal strength demonstrates a robust, statistically significant linear coupling with cortical thickness (*r* = 0.73, *p <* 0.001, Fig. 6.a). Conversely, the conventional SC nodal strength exhibits a weak and non-significant relationship with the thickness of the regional GM (*r* = 0.10, *p* = 0.35, Fig. 6.b). The high correlation with GM morphometry does not imply that DSN is redundant with cortical thickness; rather, it confirms that DSN successfully grounds complex whole-brain network topologies in valid biological tissue integrity. Unlike 1D univariate cortical thickness metrics (which are strictly local, fail to capture multi-regional network interactions [29, 44], and cannot be mathematically calculated in deep subcortical structures [45, 46]), DSN maps 2D multivariate graph topologies.

**Fig. 6.**
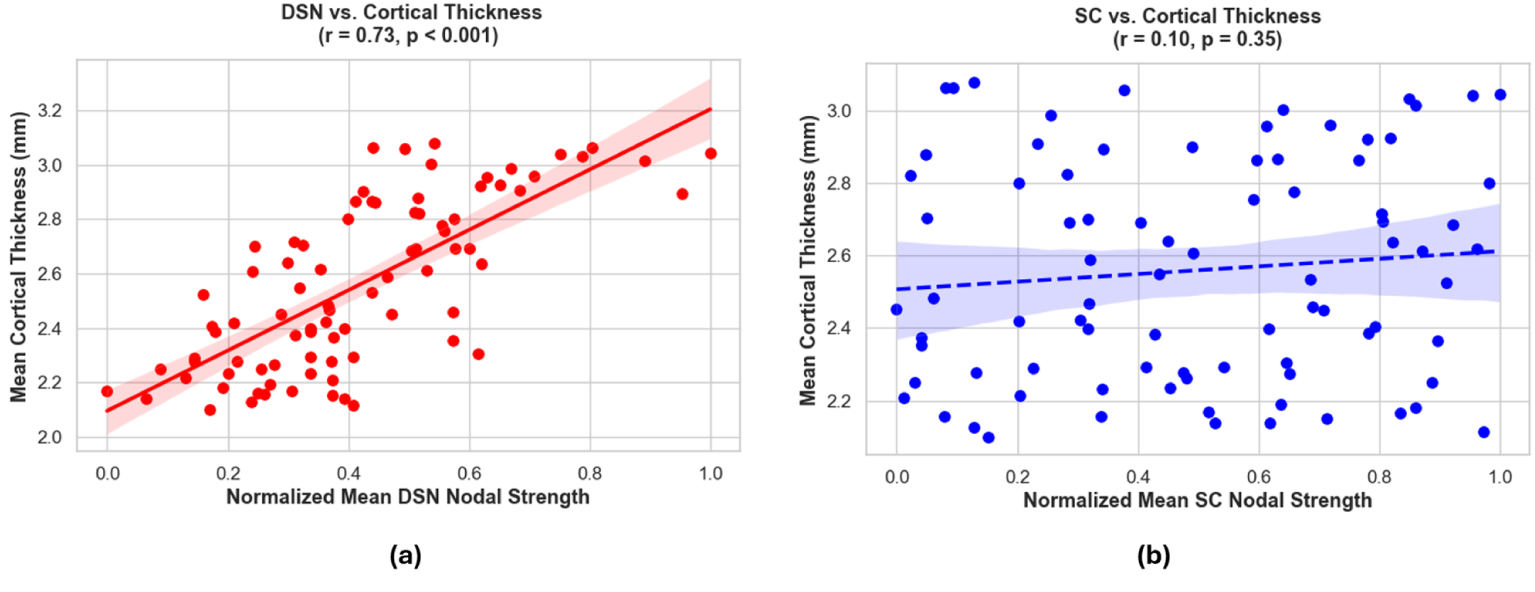
Validation of Gray-Matter microstructural alignment with (a) DSNs and (b) SC matrices.

### 4.1 Cross-Sectional DSN Validation: PD vs. Healthy Controls

To perform a baseline cross-sectional validation of the proposed DSN framework, we utilized the baseline scans from the PPMI sub-cohort, consisting of 41 individuals diagnosed with PD and 66 age-matched healthy controls (HC). The primary objective was to determine whether DSN features exhibit sufficient sensitivity to capture the microstructural distinctions between the healthy and diseased states at the group level.

First, we performed a qualitative network structure analysis utilizing Similarity Network Fusion (SNF) [31]. SNF constructs individual network similarity graphs for each subject within a group and then iteratively updates and fuses them through a non-linear method that leverages K-nearest neighbors. This integration suppresses weak, noisy edges while systematically reinforcing strong, shared topological features across the cohort, ultimately yielding a unified, representative fused group graph for both the PD and HC populations.

The resulting fused group networks are illustrated in Figure 7. A visual comparison of the two graphs reveals notable structural differences in network topology between the groups. The fused network representing the HC cohort demonstrates a highly integrated and interconnected architecture, characterized by strong cross-subnetwork communication pathways—particularly linking the default mode, limbic, and frontoparietal systems across distant anatomical clusters. In contrast, the fused group network for the PD cohort displays a markedly altered topology, where the network structure appears noticeably isolated and segmented. In the PD group, many of the long-range, cross-network edges present in the healthy controls are absent, leaving the individual functional subnetworks clustered tightly within themselves but largely disconnected from the rest of the brain. This structural isolation strongly suggests that DSNs successfully capture a widespread loss of global network integration and microstructural degeneration associated with PD cohort.

**Fig. 7.**
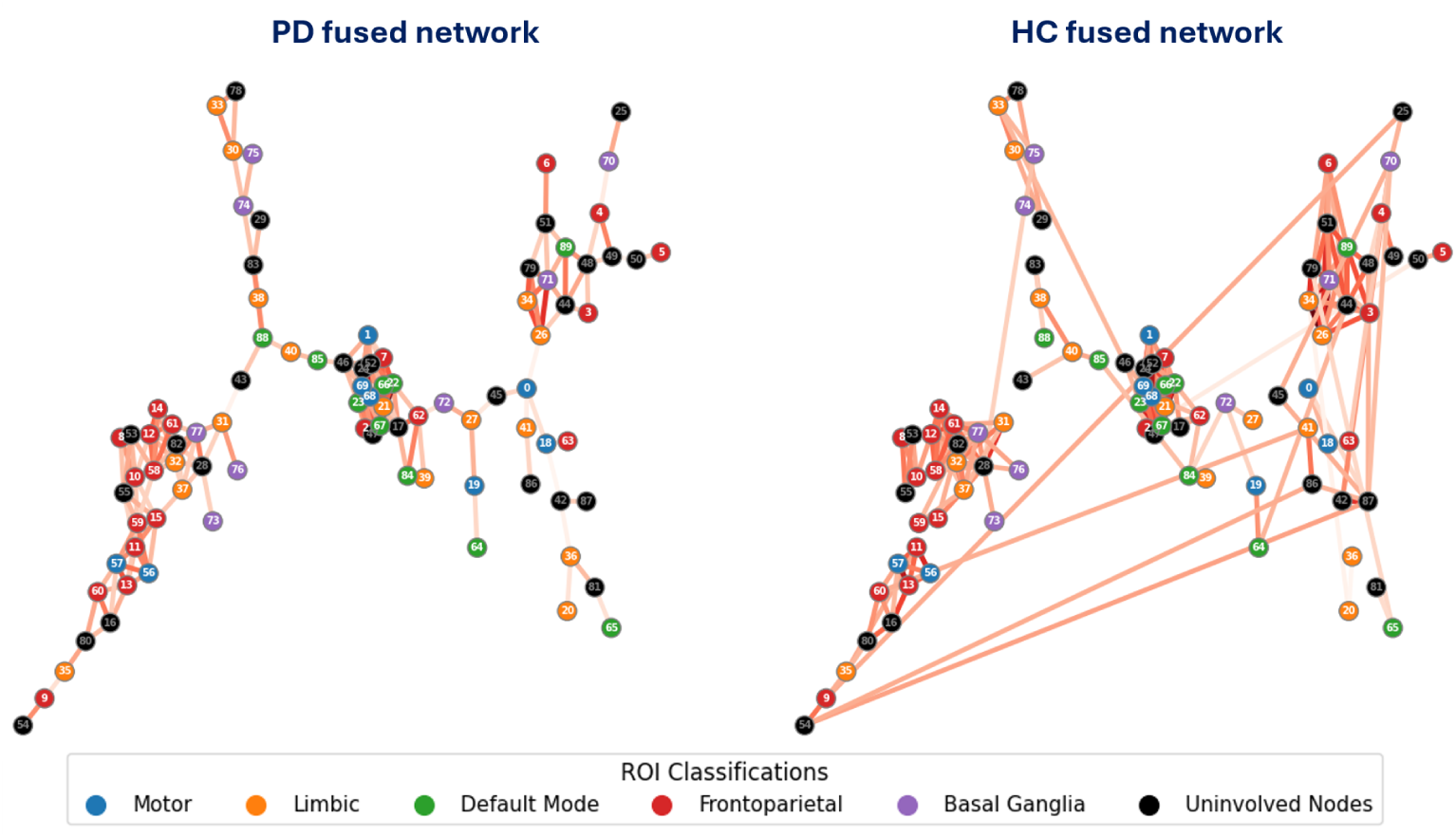
Visual comparison of fused group networks for PD patients and healthy controls generated via SNF. Nodes correspond to the 90 parcellated brain regions and are color-coded by their functional subnetwork classification (Motor, Limbic, Default Mode, Frontoparietal, Basal Ganglia, and Uninvolved Nodes.

Second, to quantify the changes in network topology suggested by the visual fusion results, we utilized the Brain Connectivity Toolbox (BCT) [47]. This toolbox enables evaluation of changes in network segregation, integration, and the structural influence of individual nodes using formal graph-theoretical metrics.

#### 4.1.1 Global Network Metrics

We first evaluated global network behavior at a standard matrix density threshold of 20% across three primary metrics: transitivity, clustering coefficient, and global efficiency (Figure 8). Transitivity and clustering coefficient serve as indicators of network segregation, reflecting the tendency of the brain to form highly localized processing clusters. Global efficiency measures network integration, representing the capacity of the connectome to rapidly transmit parallel information across long-range and distributed pathways.

**Fig. 8.**
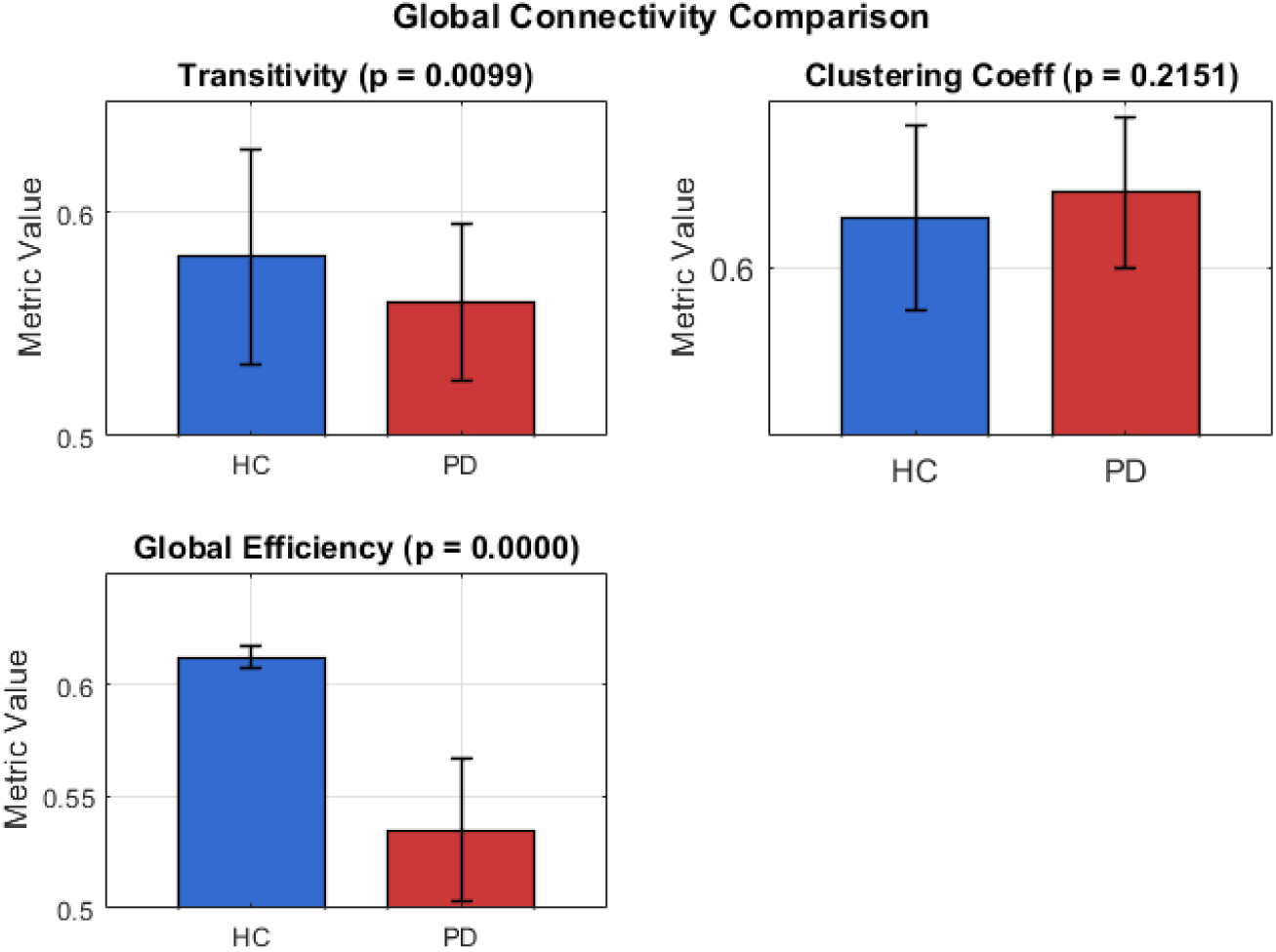
Global network topology comparison between healthy controls (HC, blue) and Parkinson’s disease patients (PD, red) evaluated at a standard matrix density threshold of 20%. Metric values represent mean *±* standard error. The PD group exhibits a significant reduction in transitivity (*p* = 0.0099) and a highly significant collapse in global efficiency (*p <* 0.0001), indicating a marked loss of long-range network integration. No statistically significant difference was observed in the mean clustering coefficient (*p* = 0.2151).

As shown in Fig. 8, a significant reduction was observed in the PD group for both transitivity (*p* = 0.0099) and global efficiency (*p <* 0.0001). The sharp drop in global efficiency mathematically underpins the loss of long-range connecting paths observed in the SNF results, demonstrating a systemic disruption in information routing efficiency across the brain. Conversely, the mean clustering coefficient did not show a statistically significant difference between groups (*p* = 0.2151). This selective preservation of local clustering, paired with a significant drop in global efficiency and transitivity, indicates that while local network modules remain relatively intact, the broader, high-efficiency topological structure needed for global cognitive and motor integration is heavily degraded in PD.

#### 4.1.2 Nodal Metrics across Functional Subnetworks

To determine exactly where these topological breakdowns occur, we mapped nodal alterations—specifically betweenness centrality and node degree differences—across target functional subnetworks (Figure 9). Betweenness centrality identifies critical hub regions by measuring how frequently a node acts as a mandatory bridge along the shortest path between all other pairs of nodes. Node degree calculates the total number of connections attached to a node, with the degree difference (Δ Connection Count) tracking net structural gains or losses in the PD group relative to the HC baseline.

**Fig. 9.**
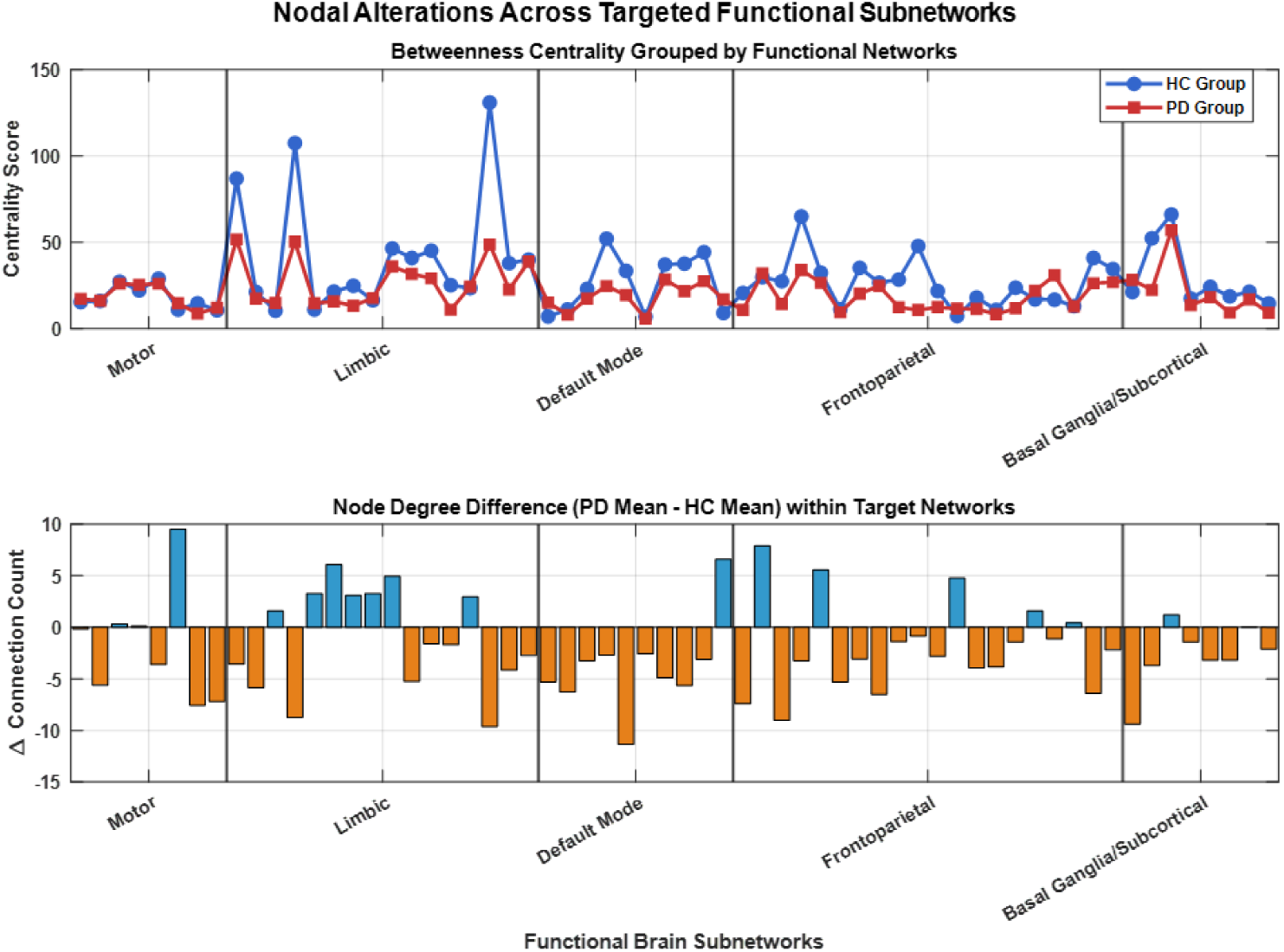
Nodal topology alterations mapped across targeted functional brain subnetworks (Motor, Limbic, Default Mode, Frontoparietal, and Basal Ganglia/Subcortical). **Top Panel:** Betweenness centrality profiles for the HC group (blue) and PD group (red), illustrating a generalized flattening of centrality peaks and a loss of hub status in the PD group. **Bottom Panel:** Node degree differences (Δ Connection Count = PD Mean *−* HC Mean) across individual network nodes. Orange bars indicate net connection losses in PD, while blue bars represent localized gains, highlighting severe structural degradation within the Default Mode Network alongside isolated compensatory increases in the Motor and Limbic networks.

The nodal results reveal severe, targeted network degradation. As illustrated in the top panel of Figure 9, the betweenness centrality scores for the PD group are depressed compared to the HC group, displaying a flattening of centrality peaks across all major subnetworks. This collapse is consistent within the Default Mode Network (DMN), Limbic system, and Frontoparietal regions.

This drop in hub status is directly explained by the node degree differences shown in the bottom panel of Figure 9. The DMN displays a massive, sustained loss in connection count (depicted by orange negative bars), reaching a deficit of over 10 connections at its most severely affected node. Widespread connection losses are also visible throughout the Motor, Limbic, and Basal Ganglia/Subcortical networks. Interestingly, a few highly localized nodes in the Motor and Limbic networks exhibit isolated increases in node degree (blue positive bars). These localized increases likely represent compensatory path remodeling, where the brain attempts to form local bypasses to offset the widespread loss of primary microstructural channels. Taken together, these BCT results demonstrate that DSNs successfully capture both global communication failures and specific subnetwork vulnerabilities in PD.

### 4.2 Longitudinal Analysis: PD Progression Trajectories

To evaluate the utility of our framework in modeling disease progression over time, we deployed a longitudinal tracking pipeline using a sub-cohort of 41 PD patients from the PPMI database. Each subject had complete imaging and clinical evaluations across four time points spanning a 4-year period (Baseline, Year 1, Year 2, and Year 4). Raw diffusion data were preprocessed and converted into CSS matrices to appropriately capture absolute structural degeneration over time without parallel degradation problem.

Pearson correlation analysis revealed significant positive correlation between the rate of microstructural network loss and the rate of clinical motor decline. The top 10 regions of interest (ROIs) exhibiting the highest correlation values are reported in Table 3, while the complete set of correlation values and corresponding graphs for all 90 anatomical ROIs are provided in the Supplementary Materials.

**Table 3.** Top 10 ROIs showing the highest correlation between microstructural brain loss and clinical motor decline.

| Anatomical Region Name (AAL3) | Pearson Correlation ($r$) | Significance ($p$) |
| --- | --- | --- |
| Supp_Motor_Area_R | +0.3445 | 0.0274 |
| Supp_Motor_Area_L | +0.3228 | 0.0395 |
| Occipital_Inf_L | +0.3156 | 0.0444 |
| Precentral_R | +0.3100 | 0.0486 |
| Precentral_L | +0.3090 | 0.0493 |
| Paracentral_Lobule_L | +0.3067 | 0.0511 |
| Occipital_Mid_L | +0.3056 | 0.0519 |
| Occipital_Inf_R | +0.3051 | 0.0524 |
| Fusiform_L | +0.3039 | 0.0534 |
| Occipital_Sup_R | +0.3016 | 0.0553 |

Based on these results, the bilateral supplementary motor areas and primary motor cortices are considered to be the most correlated regions driving the clinical motor decline. The right and left supplementary motor areas (Supp Motor Area R: *r* = +0.3445*, p* = 0.0274; Supp Motor Area L: *r* = +0.3228*, p* = 0.0395) and primary motor cortices (Precentral R: *r* = +0.3100*, p* = 0.0486; Precentral L: *r* = +0.3090*, p* = 0.0493) demonstrated statistical significance (*p <* 0.05). Additionally, the left paracentral lobule (*r* = +0.3067*, p* = 0.0511) and multiple occipital and fusiform structures demonstrated near-significant values (*p ≈* 0.05).

To evaluate whether these dynamic network markers can distinguish clinical phenotypes, the cohort was stratified based on the slope of their long-term MDS-UPDRS III progression using *K*-Nearest Neighbors (*K*-NN) clustering. This operation effectively clustered the patients into two distinct clinical phenotypes: a *Clinically Stable Class* (slow progression group characterized by a flat or minimal clinical score slope) and a *Clinical Rapid Class* (fast progression group characterized by a steep, progressive increase in motor deficits). We then evaluated the net network alteration (ΔCSS = CSS_Year 4_ *−* CSS_Baseline_) across five primary functional subnetwork systems to compare the structural outcomes of these two phenotypes (Figure 10). As illustrated in this figure, the structural trajectories diverge dramatically between the two clinical phenotypes. For the Clinically Stable Class (green bars, 19 subjects), the ΔCSS values across all five subnetworks remain positive (ΔCSS *>* 0). This net positive change indicates that stable patients maintain structural integrity or exhibit mild localized remodeling over the 4-year period.

**Fig. 10.**
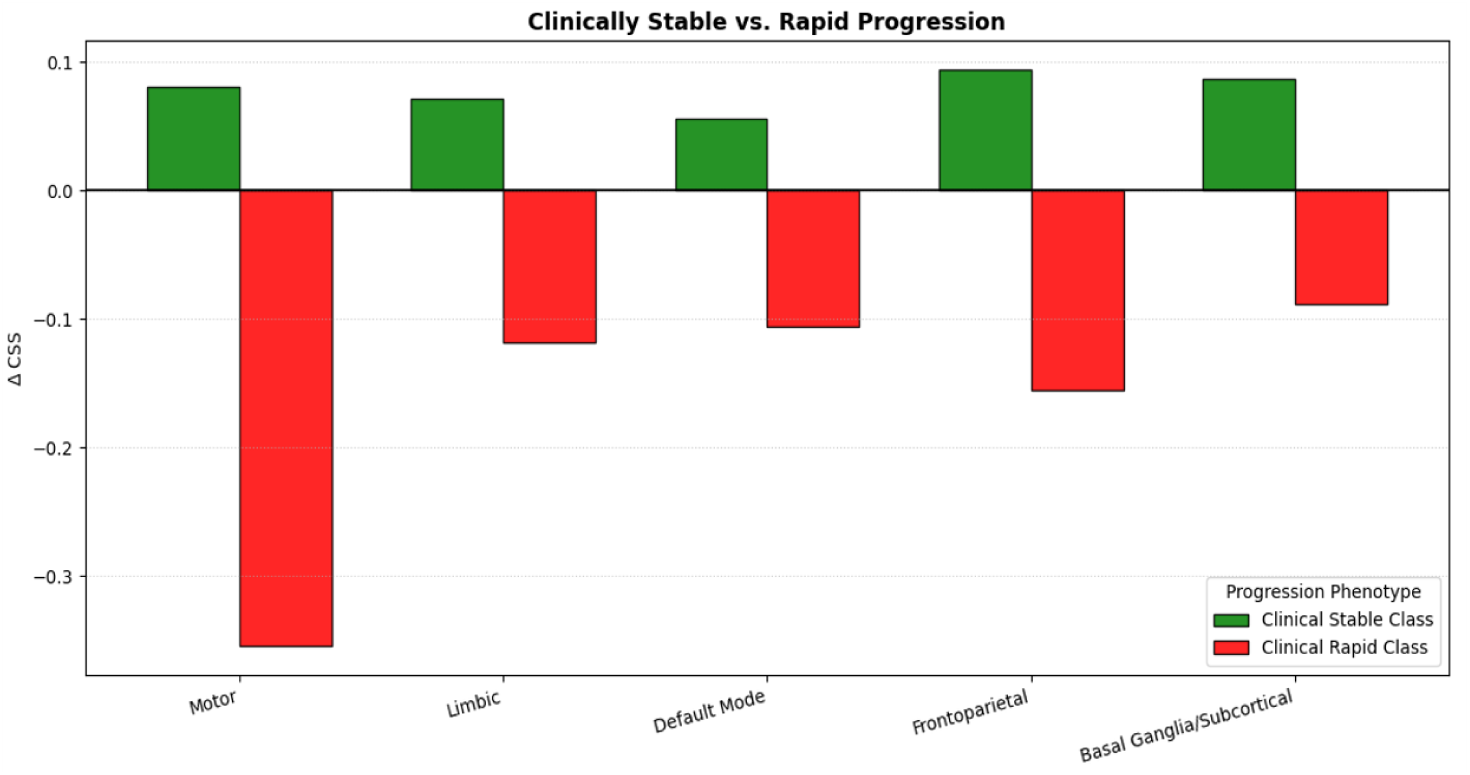
Comparison of microstructural similarity changes (ΔCSS between baseline and Year 4) across primary functional brain subnetworks for the two *K*-NN stratified phenotypes: clinically stable/slow progression class (green, 19 subjects) and clinical rapid/fast progression class (red, 22 subjects). Positive values indicate structural preservation/remodeling, while negative values indicate severe microstructural network degeneration.

On the other hand, the Clinical Rapid Class (red bars, 22 subjects) shows a progressive decline in microstructural similarity across all five examined subnetworks, with all values are negative (ΔCSS *<* 0). The most profound degeneration occurs within the Motor subnetwork (ΔCSS *≈ −*0.35), closely followed by the Frontoparietal (ΔCSS *≈ −*0.16), Limbic (ΔCSS *≈ −*0.12), and Default Mode (ΔCSS *≈ −*0.11) networks. The Basal Ganglia/Subcortical network also shows a substantial net drop (ΔCSS *≈ −*0.09).

### 4.3 Classification Model: PD vs. ET

As shown in Fig. 5, the third application establishes an advanced diagnostics classification pipeline designed to differentiate PD from ET utilizing a locally acquired cohort collected at UTSW. The clinical dataset comprises 100 total subjects balanced across classes, consisting of 57 confirmed PD patients and 43 confirmed ET patients. Validation is conducted using a stratified 5-fold cross-validation scheme. Model interpretation is achieved by GNNExplainer[48] integration, utilizing the mutual information optimization approach to generate explicit node-level salience weights mapping structural anomalies distinct to either the PD or ET clinical cohorts.

The diagnostic performance of the combined IPS-RFE and GNN framework evaluated on this dataset is summarized in Table 4. The proposed network pipeline displays strong performance, outperforming baseline models across all metrics evaluated via 5-fold cross-validation. The validation folds produced a highly stable Area Under the Receiver Operating Characteristic (ROC) curve, consistently achieving an overall AUC of 0.94.

**Table 4.** Diagnostic Performance Summary for PD vs. ET Discrimination via 5-Fold Cross-Validation.

| Model Architecture | Balanced Acc. | F1 Score | AUC |
| --- | --- | --- | --- |
| IPS-RFE + Random Forest | $0.79 \pm 0.02$ | $0.78 \pm 0.03$ | $0.81 \pm 0.02$ |
| DSN + GNN (No RFE) | $0.83 \pm 0.09$ | $0.80 \pm 0.05$ | $0.88 \pm 0.05$ |
| **DSN + IPS-RFE + GNN** | **$0.87 \pm 0.05$** | **$0.86 \pm 0.05$** | **$0.94 \pm 0.03$** |

**Fig. 11.**
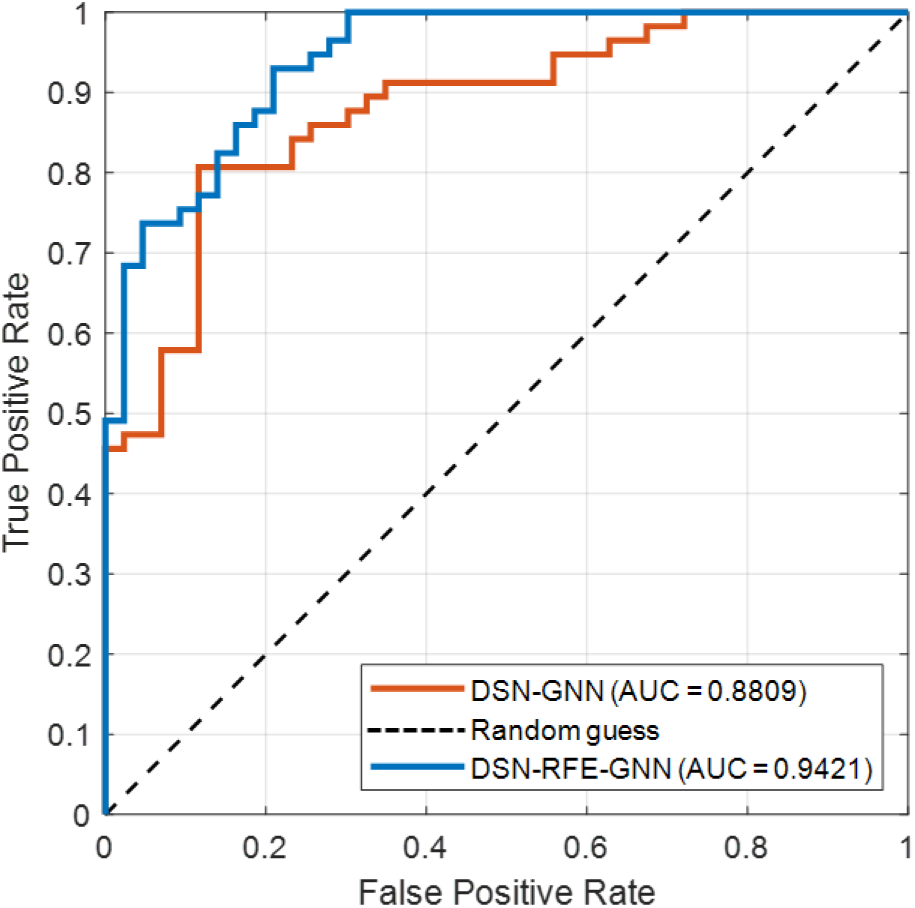
Comparison of ROC curves (with AUCs) illustrating classification performance differences between the proposed IPS-RFE GNN model with and without the IPS-RFE step.

The explainability analysis using GNNExplainer reported the top 10 structural regions that contributed most to the classification decision for each disease. Table 5 details the identified nodes ranked by relative diagnostic importance weight. This analysis indicates that PD subjects show high variation centered within striatocortical, pallidal, and mesial temporal loops. In contrast, ET structural variations are focused within the supratentorial region of the cerebello-thalamo-cortical circuit, specifically involving the thalamus, primary sensorimotor cortices, and the supplementary motor area.

**Table 5.** Top 10 Diagnostic ROIs based on the GNNExplainer importance weight.

| Rank | PD ROI | ET ROI |
| --- | --- | --- |
| 1 | Putamen (Left) | Precentral Gyrus (Right) |
| 2 | Putamen (Right) | Supplementary Motor Area (Left) |
| 3 | Caudate (Left) | Thalamus (Left) |
| 4 | Pallidum (Left) | Thalamus (Right) |
| 5 | Pallidum (Right) | Precentral Gyrus (Left) |
| 6 | Precentral Gyrus(Left) | Postcentral Gyrus(Right) |
| 7 | Supplementary Motor Area (SMA) | Superior Parietal Gyrus (Left) |
| 8 | Caudate (Right) | Supplementary Motor Area (Right) |
| 9 | Middle Frontal Gyrus (Left) | Postcentral Gyrus (Left) |
| 10 | Hippocampus (Left) | Middle Frontal Gyrus (Right) |

To confirm generalization and robustness against overfitting, the trained IPS-RFE + GNN pipeline was tested against a separate hold-out clinical validation set matching 40 total subjects (20 PD and 20 ET) locally collected at UTSW and different from the original 100 subjecys used during cross-validation. In this unseen set, the model achieved a high generalization capacity, producing a balanced testing accuracy of **87.5%**.

## 5 Discussion

Characterizing individual structural alterations in movement disorders has remained a long-standing challenge due to the inability of standard structural imaging or white matter tractography to fully capture subtle, localized gray matter microstructural changes. In this study, we addressed this limitation by developing and validating a unified framework centered on DSNs. By modeling the brain as a graph built on microstructural similarities rather than raw fiber counts, our findings demonstrate that DSNs provide a highly sensitive, clinically relevant biomarker for studying disease pathology, tracking progression, and improving diagnostic accuracy.

Our cross-sectional validation confirmed that DSNs possess the sensitivity required to distinguish diseased states from healthy controls. The qualitative network topographies generated via SNF highlighted a clear structural divergence: while healthy control networks maintain high global integration, the PD networks exhibit pronounced segmentation, with functional subsystems collapsing into isolated clusters.

This visual isolation was robustly quantified using the BCT. The significant reduction in global efficiency and transitivity within the PD cohort mathematically confirms a systemic breakdown in long-range communication pathways. Interestingly, the preservation of the local clustering coefficient alongside severe connection count drops—particularly within the Default Mode Network —suggests that PD topology is characterized by a selective failure of long-range integrative hubs rather than a uniform destruction of local modules.

A primary innovation of this work is the adaptation of the DSN framework into the CSS metric for longitudinal modeling. Standard similarity metrics suffer from a problem where parallel degradation in two communicating regions yields an artificially stable correlation over time. By incorporating an exponential decay penalty based on absolute tissue status, the CSS metric successfully unmasked true longitudinal decline. Our longitudinal velocity coupling framework revealed that the velocity of CSS network decay significantly correlates with the accumulation rate of clinical motor symptoms (MDS-UPDRS III). The fact that the bilateral supplementary motor areas and primary motor cortices emerged as the most highly correlated regions provides profound neurobiological validation, as these exact regions serve as the structural epicenters for motor planning and execution. Furthermore, using K-NN clustering to stratify patients based on clinical trajectory showed that rapid progressors experience a severe, widespread network collapse across all primary subnetwork systems (dominated by the motor network), whereas stable/slow progression patients maintain structural integrity or exhibit localized compensatory remodeling over a four-year period. These results highlight the potential of longitudinal CSS mapping to serve as a predictive biomarker for patient stratification in clinical trials.

Finally, the translation of these microstructural networks into a predictive GNN framework demonstrated excellent diagnostic accuracy. By deploying our IPS-RFE method, we successfully compressed the high-dimensional feature space of 4,005 connections down to the top 15% highly discriminant connections. Using these filtered DSN topologies as graph embeddings allowed the GNN to effectively navigate the challenging differential diagnosis task between PD and ET within our local UTSW cohort. This confirms that DSN features contain highly specific, disease-defining signatures capable of separating distinct movement disorder phenotypes that can present with similar clinical symptoms.

This study provides several key contributions to the field of clinical neuroscience and computational neuroimaging:

- **Methodological Innovation (CSS Metric):** We introduced the CSS metric, which overcomes the limitations of traditional similarity matrices in longitudinal studies by penalizing connections based on absolute regional degradation, successfully capturing true structural decay over time.
- **Longitudinal Velocity Coupling:** We established a novel workflow to map imaging metrics and clinical trajectories into a unified 2D velocity coordinate space, directly demonstrating that faster microstructural network decay drives faster clinical motor decline.
- **High-Dimensional Diagnostic Performance:** By pairing DSN inputs with an IPS-RFE feature selection routine and a GNN architecture, we established a robust and accurate framework for the differential diagnosis of complex, overlapping movement disorders (PD vs. ET).

Despite the promising performance and validation of the framework, several limitations must be acknowledged. First, the longitudinal analysis was restricted to a highly curated sub-cohort from the PPMI database to ensure perfectly age-matched controls and complete four-session timelines over four years. While this provided excellent internal consistency, it reduced the available sample size for progression modeling. Future work will focus on validating these longitudinal velocity models on larger, more heterogeneous datasets.

Second, the structural degradation metrics used to penalize the CSS equation were derived based on deviations from a group-level healthy baseline. While effective, developing a fully individualized status index that tracks baseline deviations relative to a patient’s own initial scan could further personalize the metric’s sensitivity. Finally, while the GNN accurately classified PD from ET on our local UTSW cohort, expanding this framework into a multi-class deep learning model that includes atypical Parkinsonian variants (such as Multiple System Atrophy or Progressive Supranuclear Palsy) would represent a critical step toward true clinical utility in movement disorder clinics. Furthermore, these results would benefit from validation in an external cohort.

## 6 Conclusion

In conclusion, this study demonstrates that DSNs provide a robust, highly sensitive, and comprehensive framework for characterizing microstructural brain network alterations in movement disorders. By mapping individual brain regions as similarity nodes, our pipeline successfully captured global communication failures and localized subnetwork breakdowns in Parkinson’s disease. The introduction of the CSS metric and the longitudinal velocity coupling framework successfully bridged the gap between MRI and clinical motor decline, providing an quantifiable biomarker to track and stratify disease progression phenotypes over time. Furthermore, combining these refined networks with a GNN architecture demonstrated powerful diagnostic utility in differentiating PD from ET. Taken together, these findings highlight the potential of diffusion similarity mapping to serve as a powerful computational biomarker, offering detailed insights into neurodegenerative progression and improving clinical management.

## Data Availability

All data produced in the present study are available upon reasonable request to the authors

